# SARS-CoV-2 viral load in the upper respiratory tract of children and adults with early acute COVID-19

**DOI:** 10.1101/2020.07.17.20155333

**Authors:** Stéphanie Baggio, Arnaud G. L’Huillier, Sabine Yerly, Mathilde Bellon, Noémie Wagner, Marie Rohr, Angela Huttner, Géraldine Blanchard-Rohner, Natasha Loevy, Laurent Kaiser, Frédérique Jacquerioz, Isabella Eckerle

**Affiliations:** Division of Prison Health, Geneva University Hospitals, Geneva, Switzerland; Office of Corrections, Department of Justice and Home Affairs of the Canton of Zurich, Zurich, Switzerland; Children’s Hospital of Geneva, Geneva University Hospitals and Faculty of Medicine, Geneva, Switzerland; Laboratory of Virology, Division of Laboratory Medicine, Geneva University Hospitals and Faculty of Medicine, Geneva, Switzerland; Division of Infectious Diseases, Department of Medicine, Geneva University Hospitals and Faculty of Medicine, Geneva, Switzerland; Center for Vaccinology and Department of Pediatrics, University Hospitals of Geneva, Geneva, Switzerland; Pediatric Platform for Clinical Research, Department of Woman, Child and Adolescent Medicine, Geneva University Hospitals and Faculty of Medicine, University of Geneva, Geneva, Switzerland; Center for Emerging Viral Diseases, Geneva University Hospitals and Faculty of Medicine, Geneva, Switzerland; Division of Tropical and Humanitarian Medicine, Geneva University Hospitals and Faculty of Medicine, Geneva, Switzerland; Primary Care Division, Geneva University Hospitals and Faculty of Medicine, Geneva, Switzerland; Department of Molecular Medicine and Microbiology, Faculty of Medicine, Université de Genève, Geneva, Switzerland

## Abstract

The role of children in the transmission of SARS-CoV-2 is unclear. We analysed viral load at the time of diagnosis in 53 children vs. 352 adults with COVID-19 in the first 5 days post symptom onset. No significant differences in SARS-CoV-2 RNA loads were seen between children and adults.

## Introduction

Children are underrepresented in the current SARS-CoV-2 pandemic, and most infected children exhibit limited disease severity or are asymptomatic [1]. Although SARS-CoV-2 infects children of all ages, children do not seem to be major drivers of transmission, unlike for other respiratory viruses [2–4]. Current epidemiological data suggest a lower susceptibility among children, although most studies were performed while lock-downs/infection-prevention measures were in place, which included school closures [5]. Also, so far most documented pediatric infections originate from an adult case [2–4]. Furthermore, the role of children in spreading the virus to others is not well understood; in this regard, the kinetics and peak viral loads (VL), key determinants of virus transmission, have to be compared with VL of adults. Recent SARS-CoV-2 RNA load comparisons across age groups have yielded inconsistent conclusions, finding similar or lower VL in children depending on the type of statistical analysis used, sample-collection time period, and diagnostic assay used [6–8]. Further understanding of VLs across age groups is relevant, as multiple studies established thresholds for the presence of infectious SARS-CoV-2, as assessed by isolation of culture-competent SARS-CoV-2 on cell lines [9–12]. Thus, viral RNA load can also serve as a surrogate of infectious virus shedding and potential transmission capacity.

In this single-centre study, we investigated SARS-CoV-2 RNA load from the URT at the time of diagnosis among symptomatic children and adults consulting our hospital in their first 5 days post onset of symptoms (DPOS).

## Methods

### Study design and setting

Data were collected in the context of prospective cohort studies at a single-centre. All diagnostic testing was performed at the virology laboratory of the Geneva University Hospitals (HUG), a tertiary-care centre that serves the canton of Geneva and also serves as the Swiss National Reference Centre for Emerging Viral Diseases. Data on SARS-CoV-2 infected outpatients ≥16 years old were collected at HUG’s SARS-CoV-2 testing centre, and data on patients <16 years old were collected for all children who tested positive for SARS-CoV-2 in our laboratory.

### Participants

Participants were included if they had a positive SARS-CoV-2 RT-PCR but excluded if 1) a previous positive SARS-CoV-2 test, 2) more than five DPOS 3) no symptoms at the time of diagnosis.

### Measures

Diagnostic specimens consisted of nasopharyngeal swab samples collected with a flocked swab and stored in viral transport medium. Specimens in the presented analysis were either tested by an in-house method based on the Charité E gene assay or the Cobas 6800 SARS CoV2 RT-PCR (Roche, Switzerland), targeting the E and the ORF1 gene, which were the two main diagnostic methods used in our Centre and showed a close correlation in terms of sensitivity and specifity [13]. A standard curve was obtained for both systems by quantifying *in vitro* transcribed RNA (in-house method) or quantified supernatant from a cell culture isolate of SARS-CoV-2 (Cobas) [10]. For both assays, a formula for calculation of VLs was obtained and SARS-CoV-2 RNA copy numbers from all samples were calculated from the cycle threshold values (CT values), in detail this was (CT-44.5)/-3.3372 for Cobas (E gene) and (CT-41.7)/-3.4529 for the in-house assay (E gene). As VL was skewed, we used a logarithmic transformation (log10).

### Age

Age was collected at the day of the visit. The following categories were assigned: child (0-11), adolescent (12-19), young adult (20-45), and adult (>45 years).

### Statistical analyses

We calculated the sample size based on a correlation coefficient using z transformation[14]; with alpha=.05, power=.80, Pearson r=.20, the required sample size was n=194. We first computed descriptive statistics for all variables. We then tested the association between age and VL. We plotted age (continuous) against log10 VL and computed Pearson’s correlation. Finally, we tested whether age groups were associated with VL using one-way analysis of variance, using *post-hoc* pairwise comparisons with Bonferroni adjustment between age groups and polynomial contrasts to identify trends. All analyses were performed with Stata 15. The sample size calculation was estimated using www2.ccrb.cuhk.edu.hk/stat.

### Ethics

The work was approved by the Cantonal ethics committee (no. 2020-00813; no. 2020-00835, no. 2020-00516). All study participants and/or their legal guardians provided written informed consent.

## Results

For patients ≥16 years old, we identified 352 patients fulfilling our inclusion criteria, that tested positive for SARS-CoV-2 in a selected time period between March 29 and April 23, 2020. For patients < 16 years old, we identified 53 children fulfilling our inclusion criteria that tested positive for SARS-CoV-2 in a selected testing period between March 10, 2020 and 26 May, 2020. The total sample size was thus 405.

Mean age of all patients was 36.53 ± 16.26 years (range 0-82); mean log10 VL was 6.06 ± 1.98 RNA copies/mL (range 1.52-9.42). Children and adolescents had a mean average VL of 6.13 ± 2.02 (range 3.06-9.21) and 5.85 ± 2.32 (range 2.36-9.42) log10 RNA copies/mL, respectively, versus the two adult groups’ (20-45 and >46 years) means of 5.91 ± 1.88 (range 2.37-9.39) and 6.33 ± 2.05 (range 2.49-9.39), respectively. VLs that lie above a postulated threshold for infectious virus shedding of around 6 log10 RNA copies was reached for 49.5 – 62.6 % of patients (Supplementary table S1).

The correlation between age (continuous) and log10 VL was *r*=.01 (*p*=.797), as shown in Figure 1A. In the one-way analysis of variance, the association of age (categories) with VL was not significant (*F_3, 401_*=1.34, *p*=.259). Neither *post-hoc* pairwise comparisons nor polynomial contrasts were significant (Bonferroni corrected p-values≥.333; linear contrast: *F_1, 401_*=0.29, *p*=.591; quadratic contrast: *F_1, 401_*=1.54, *p*=.216). Figure 1B shows the relationship between age groups and log10 VL.

**Figure 1.**
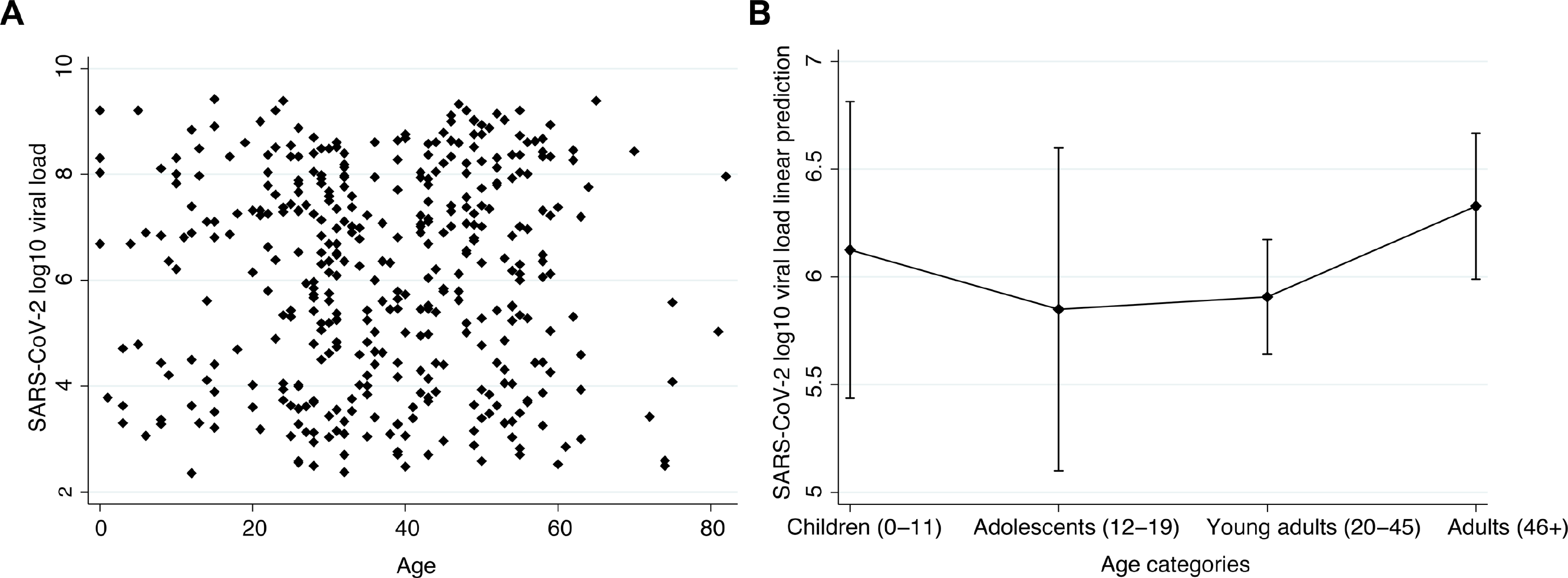
A. Scatterplot of age (continuous) against log10 viral load B. Means and 95% confidence intervals of log10 viral load against age groups.

## Discussion

We compared VL among different age groups to clarify the role of children in SARS-CoV-2 transmission. Our main finding was that VL was similar across all age ranges, using age as a continuous variable and as a categorical factor.

In studies quantifying VL, observed thresholds for successful isolation were in the range of 5.4 - 7 log10 RNA copies/mL, or a CT value of 24 and below for another study using only the test-specific CT for their correlation, which is expected to correlate with the same copy number range [9–12]. Here we could show that around half or more of all samples from both children and adults have a VL above such a threshold. In addition, we recently showed successful SARS-CoV-2 isolation from 12 of 23 children with COVID-19, with a median of 7.2 log10 RNA copies/mL in culture competent samples [10].

A contested preprint of a study comparing viral loads of 3303 patients in Germany initially concluded that VL were similar across age groups, while a separate analysis of the same data set reported VL increased with age [6, 8]. In a revised version of the original manuscript, similar VL across age groups were found in a setting at the beginning of the pandemic when mainly community and cluster screening was done [7]. Slightly lower VL in children was reported at a later stage when tested with another assay and when household testing was reduced. The above-mentioned study however lacks information on symptom onset date, and shows a large heterogeneity of study subjects included in the analysis (individuals tested as part of public health investigations but also patients). Thus potential inclusion of asymptomatic contacts and patients in later infection stages with declining VLs could be confounders. In our study, we included only patients presenting within the first 5 DPOS, as it was inferred that infectiousness peaks on or before symptom onset [15], with infectious virus presence largely limited to the first week of illness. Our findings, focusing on symptomatic patients during the early phase of disease, support an absence of difference in VL among children and adults in the first 5 days. One limitation of our study was a limited sample size of children, thus age group comparisons might have lacked power. Another limitation was inclusion of symptomatic patients only, thus not reflecting the whole spectrum of SARS-CoV-2 infection, especially the asymptomatic presentation frequently seen in children. Furthermore, we included patients tested by two different methods, however both of which showed close correlation for sensitivity, specificity and for VL quantification by an external standard curve.

To our knowledge, our study is the largest analysis on VL across age groups for which time of symptom onset is available, and here analysing age as a continuous variable clearly showed no relationship between VL and age. As we found similar VL in children versus adults and around half of specimens above a threshold correlated with infectious virus presence, our results suggest that the presumed lower transmission from children is not due to lower VL in the URT compared to adults. As VL may not be the only factor for effective SARS-CoV-2 transmission, other factors might contribute to less efficient transmission in children, even with high VLs. Further studies are needed to understand the role of children in transmission, especially now that containment measures are lifted in most countries, to enable children to safely benefit from education and much needed social interactions among their peers.

## Data Availability

na

## Acknowledgments

We thank Claire-Anne Siegrist and Arnaud Didierlaurent for support of the study, Benjamin Meyer for help with data analysis and Erik Boehm for proof reading.

## Supplementary files

**Supplementary table 1.** Descriptive statistics

| Age categories | % (n) | Log10 viral load<sup>1</sup> | Log10 viral load > 6<sup>2</sup> | Symptoms onset (days)<sup>1</sup> |
| --- | --- | --- | --- | --- |
| Overall | 100 (405) | 6.06 (1.98, 2.36-9.42) | 54.1 (219) | 2.31 (1.41) |
| Children (0-11) | 7.9 (32) | 6.13 (2.02, 3.06-9.21) | 62.5 (20) | 1.75 (1.08) |
| Adolescents (12-19) | 6.7 (27) | 5.85 (2.32, 2.36-9.42) | 51.9 (14) | 2.30 (1.46) |
| Young adults (20-45) | 52.8 (214) | 5.91 (1.88, 2.37-9.39) | 49.5 (106) | 2.36 (1.45) |
| Adults (46+) | 32.6 (132) | 6.33 (2.05, 2.49-9.39) | 59.9 (79) | 2.38 (1.38) |

<sup>1</sup> Means, standard deviations, and range are reported.
<sup>2</sup> Percentages and n are reported.

